# Development and Internal Validation of a Field-Based Triage Tool for Lumbopelvic-Hip Dysfunction in Collegiate Athletes

**DOI:** 10.64898/2026.04.23.26351566

**Authors:** Hung-Chun Huang, Pei-Hsi Chou, Kung-Che Lee, I-Hua Chu, Ing-Jer Huang, Jing-Min Liang, Wen-Lan Wu

## Abstract

This cross-sectional derivation and internal validation study aimed to develop and internally validate a clinical triage scoring system (CTSS) for field-based identification of collegiate athletes requiring priority intervention for lumbopelvic-hip (LPH) dysfunction. A total of 864 collegiate athletes (mean age 21.3 ± 2.4 years; 80.8% male) were recruited from 10 universities. Participants underwent standardized assessments including demographic characteristics, clinical history, and LPH functional testing. Using an expert-adjudicated binary reference standard (priority intervention vs self-management), a multivariable logistic regression model was developed to derive the weighted CTSS. Model performance was evaluated using discrimination, calibration, and decision curve analysis (DCA), and internal validation was performed using 1,000 bootstrap resamples. Of the 864 participants, 463 athletes (53.6%) were classified as requiring priority intervention. The final 14-factor CTSS comprised 12 positive-weight predictors, such as localized LPH pain, muscle weakness, and higher body mass index, and 2 negative-weight predictors, positive Lasegue’s sign and hamstring weakness, which functioned served as safety-related modifiers. The model demonstrated acceptable discrimination (AUROC = 0.851, 95% CI: 0.824–0.876), with minimal optimism (optimism-corrected AUROC = 0.842) and excellent calibration (calibration slope = 1.000; calibration intercept = 0.000). A total score of ≥9 was identified as the optimal threshold, yielding a sensitivity of 84.4% and specificity of 71.8%. DCA showed greater net benefit than treat-all and treat-none strategies across clinically relevant threshold probabilities (20%–50%), with a net benefit of 0.319 at a 50% threshold probability. The CTSS may provide a pragmatic field-based triage tool to support early identification of athletes who may require priority intervention, although external validation is needed before broader implementation in sports medicine settings.

## Introduction

The lumbopelvic-hip (LPH) complex plays an important role in force transfer between the trunk and lower extremities during athletic movement. Impairments in this region, including muscle imbalances, joint mobility restrictions, and altered neuromuscular control, have been associated with reduced movement efficiency and may contribute to musculoskeletal injury risk in athletic populations (1, 2). Low back pain (LBP) is one of the most common clinical manifestations associated with LPH dysfunction and remains highly prevalent among athletes exposed to repetitive loading demands (1). Importantly, a history of LBP remains a consistent predictor of recurrence (1, 3), suggesting that symptomatic resolution does not necessarily reflect full restoration of neuromuscular function. Persistent alterations, such as impaired trunk muscle activation and increased spinal stiffness, have also been reported in asymptomatic athletes, potentially contributing to ongoing mechanical vulnerability (4-6).

In multidisciplinary sports medicine settings, athletes with LPH dysfunction may be managed through priority clinical intervention, activity modification, or self-management depending on clinical presentation (7). In sideline and competition settings, sports medicine practitioners, including sports chiropractors, physiotherapists, and athletic trainers, are often required to rapidly distinguish athletes who require priority intervention from those suitable for self-management. However, many assessment approaches rely on imaging, specialist evaluation, or clinic-based resources, which may be impractical in time-constrained field environments (8). As a result, triage decisions in these settings are often based on clinician judgment rather than a standardized framework.

Although clinical prediction rules have been developed for identifying candidates for specific interventions in general LBP populations (9, 10), their applicability to athletic field-based triage remains limited. Furthermore, while traditional single-item screening tests have been criticized for their limited predictive value in sports medicine (11), tools developed for established pain syndromes may be less suited to the dynamic functional impairments characteristic of high-load athletic participation. Although various functional screening instruments have been described, few have undergone rigorous internal validation, and even fewer have been explicitly linked to structured triage pathways in athletic populations (8). This gap is particularly relevant in collegiate sports settings. Therefore, this study aimed to develop and internally validate a field-applicable Clinical Triage Scoring System (CTSS) for identifying collegiate athletes requiring priority intervention for LPH dysfunction. The CTSS was developed as a decision-support tool for triage and care prioritization, rather than as a diagnostic instrument. By integrating clinical history with objective functional indicators, including range of motion, muscle performance, and pain distribution, this study sought to derive a weighted scoring framework to support structured and practical triage in collegiate sports medicine settings.

## Methods

### Participants and Study Design

This cross-sectional study was conducted to develop and internally validate a CTSS and was carried out between August 2021 and December 2024. This study was reported in accordance with the STROBE (Strengthening the Reporting of Observational Studies in Epidemiology) Statement (12). A total of 945 collegiate athletes were recruited from 10 universities in Taiwan, where standardized on-site assessments were performed by a trained sports medicine research team. The participant flow is detailed in Figure 1. Inclusion criteria were: (1) active roster status on a varsity team; (2) age 18–30 years; and (3) a minimum training volume of 10 hours per week. Recruitment was conducted regardless of current symptom status in order to capture a broad spectrum of functional presentations, ranging from asymptomatic to symptomatic, within the collegiate athletic population. Exclusion criteria were defined to ensure participant safety and data validity. Athletes were excluded if they presented with: (1) “Red flags” indicative of serious pathology (e.g., suspected fracture, infection, or malignancy) (13), as determined by a supervising medical doctor; (2) current acute lower-extremity injuries that physically precluded the completion of functional assessments (i.e., unable to bear weight or perform dynamic tasks due to pain/instability); (3) history of lower-extremity surgery within the past year; or (4) incomplete data on primary outcome measures. To assess the impact of prior trauma, athletes with a history of recent minor injuries (e.g., Grade I/II sprains) were retained in the study, provided they had returned to full participation and could complete all protocols without pain exacerbation. Of the initial cohort (N = 945), 81 athletes were excluded based on these criteria (20 clinical contraindications: 8 red flags, 12 acute injuries; 61 missing data), yielding a final sample of 864 participants. This study was conducted and reported in adherence to the STROBE guidelines to ensure the transparency and reliability of the prediction model (12). All participants provided written informed consent, and the study was approved by the Kaohsiung Medical University Institutional Review Board (KMUHIRB-F(II)-20210113, 2021/6/29-2026/8/31) in accordance with the Declaration of Helsinki.

**Figure 1.**
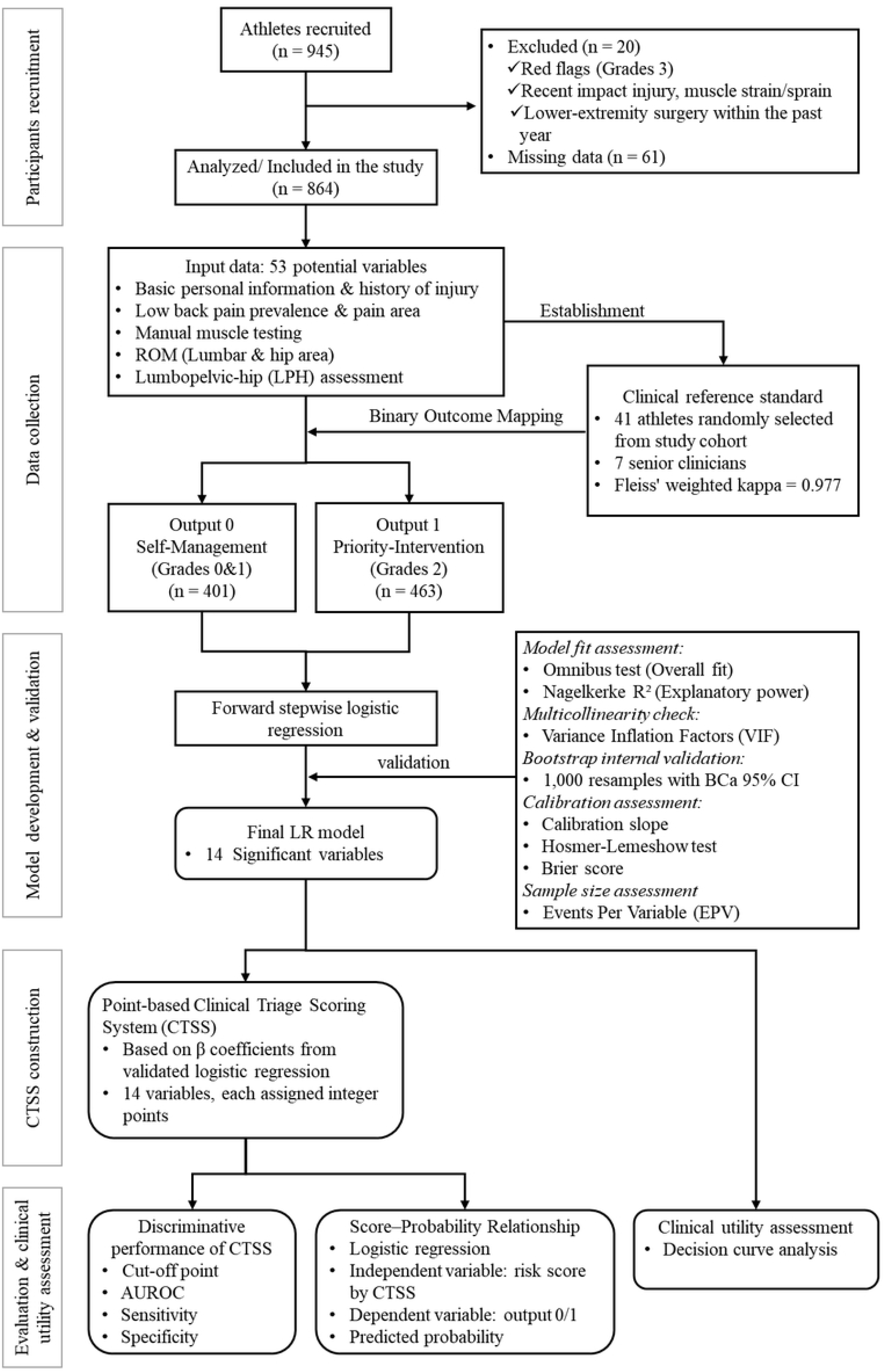
Study Flowchart

### Data Collection

Fifty-three candidate predictor variables were prespecified, guided by clinical rationale and existing literature, and categorized into four domains: (1) demographics and training characteristics; (2) retrospective injury history; (3) self-reported pain and function; and (4) objective physical examination findings. To assess these variables, a standardized, station-based assessment protocol was implemented. Examiners performing the physical assessments were blinded to participants’ demographic information, injury history, and questionnaire responses at the time of testing. The primary clinical outcome (priority intervention vs. self-management) was subsequently adjudicated independently by an expert panel after completion of data collection, thereby reducing the potential for incorporation bias.

### Demographic and Injury History

Data included sex (male/female), age (years), height (cm), weight (kg), body mass index (BMI, kg/m^2^), years of sport participation, and average weekly training volume (hours/week). The point prevalence of LBP, 6-month, and 12-month prevalence were recorded. A history of sport-related musculoskeletal injury was recorded by anatomical region (low back, pelvis, hip, knee, and ankle). Detailed variable definitions and response formats are presented in Table 1.

**Table 1.**
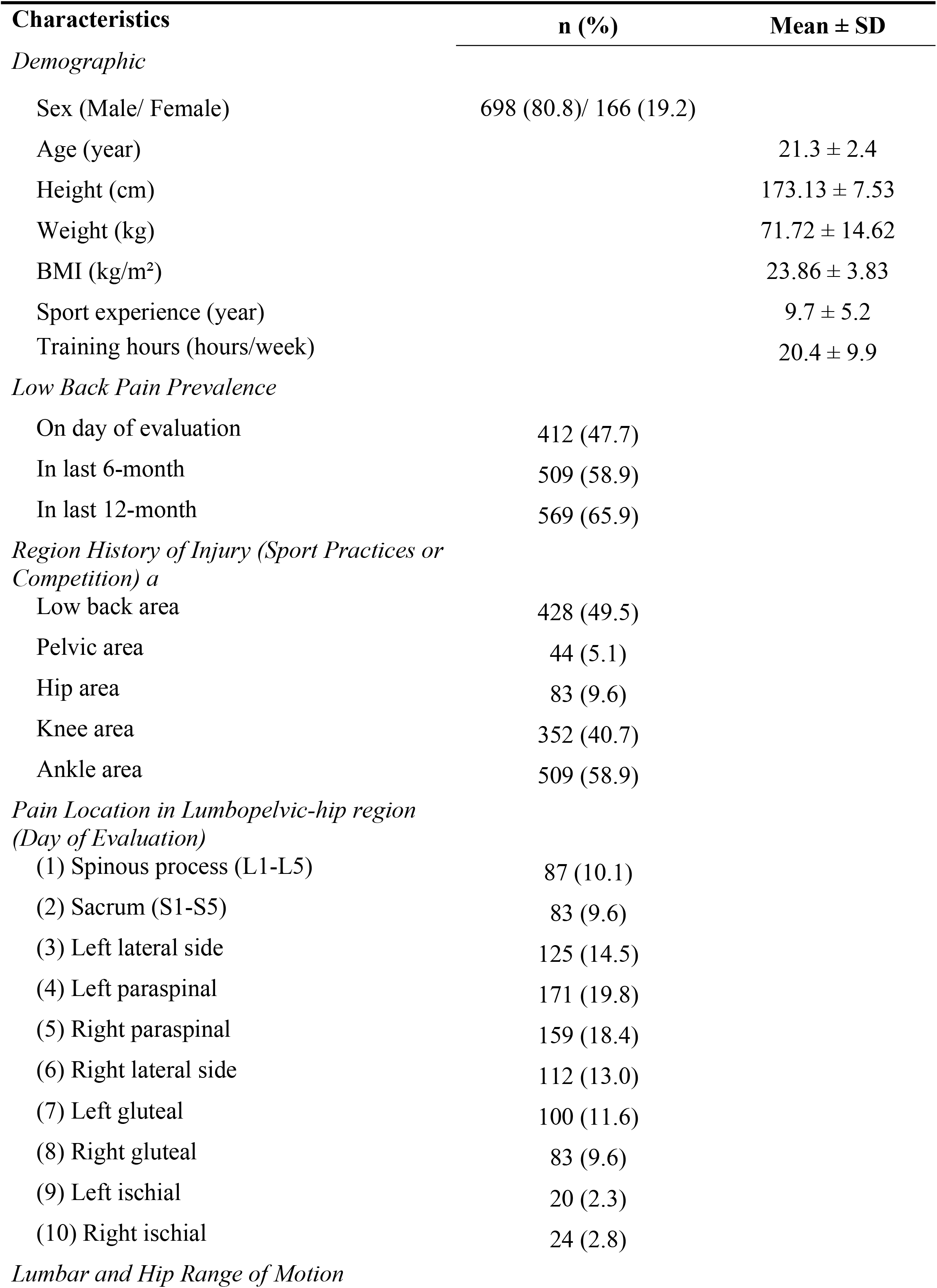
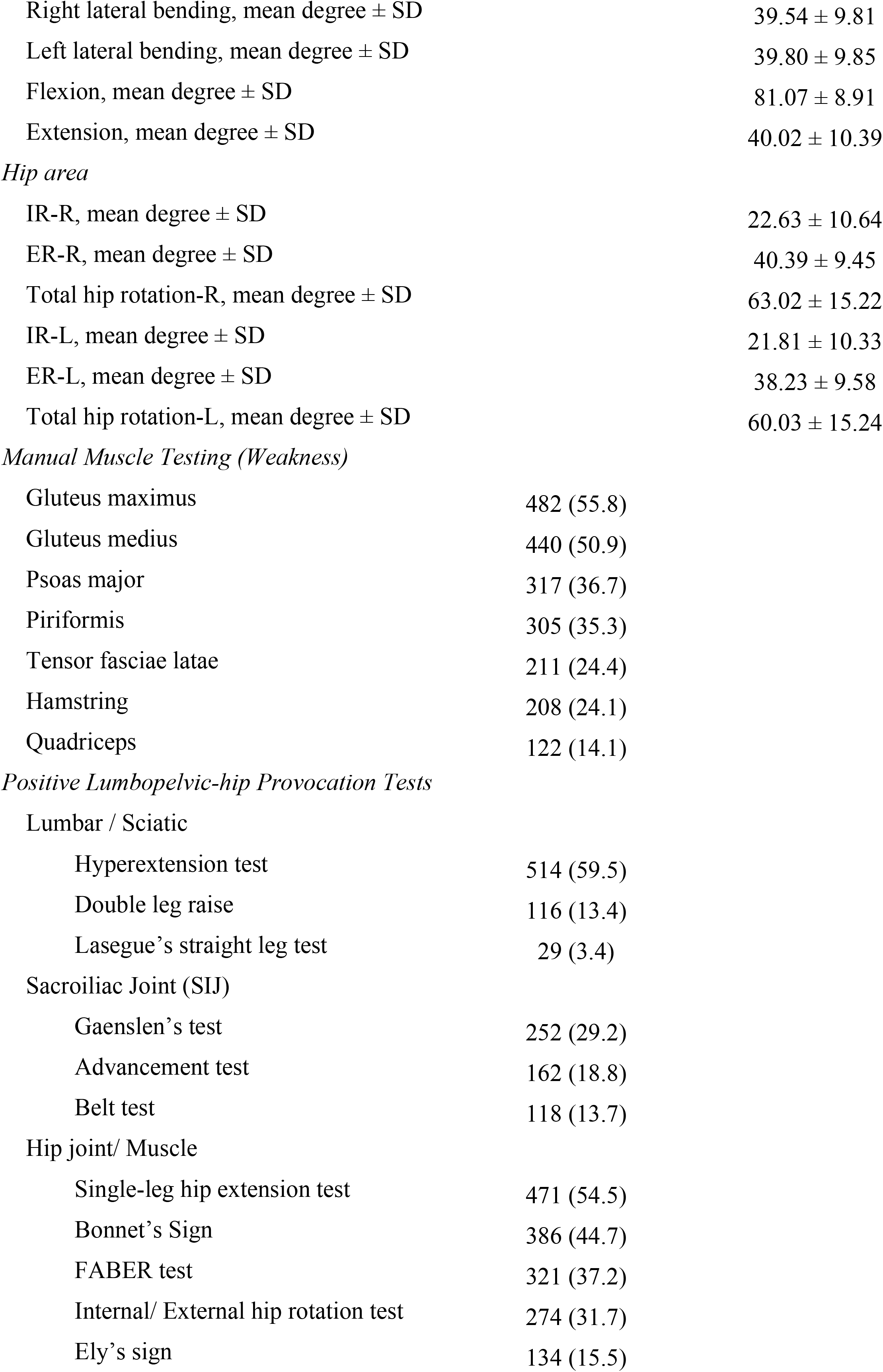
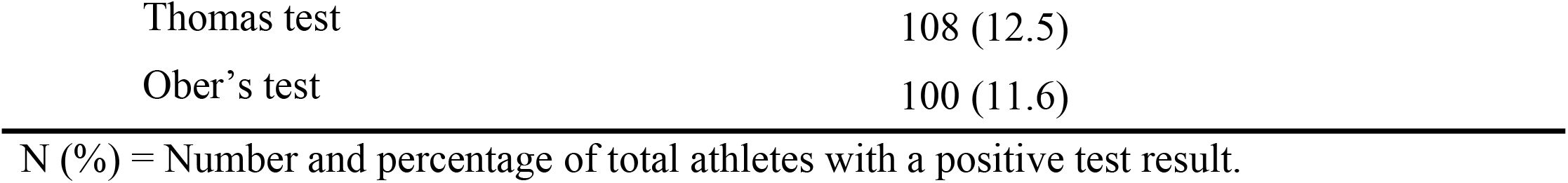
Demographic Characteristics and Prevalence of Positive Screening Findings.

### Low Back Pain Prevalence & Pain Area

LBP was anatomically defined as the region bounded superiorly by the 12th thoracic rib, inferiorly by the inferior gluteal folds, and laterally by the posterior axillary lines. Symptom localization within the LPH region was documented using a standardized 10-zone anatomical mapping system (Supplementary Figure S1), consistent with established epidemiological protocols (14).

### LBP Case Definitions

Current LBP was defined as pain or discomfort localized between the costal margin and the gluteal folds, self-reported at the time of assessment (14). A history of LBP (6- and 12-month) was defined as an episode resulting in functional consequences, including reduced training volume, impaired performance, or medical consultation, consistent with IOC consensus criteria (15, 16).

### Specific Assessment Procedures

Functional assessments were conducted using a standardized, station-based protocol adapted from established methodologies (17, 18). To minimize measurement error, all examiners completed a pre-study calibration workshop focusing on procedural standardization and scoring criteria. Testing was stratified by expertise: biomechanical measurements were led by a senior examiner (physical therapist or chiropractor), whereas all symptom-provocation tests were performed by a single experienced clinician (Doctor of Chiropractic with sports specialty certification) to eliminate inter-rater variability in symptom reproduction. A rigorous safety protocol was implemented under physician supervision, including a structured referral pathway for adverse events. All assessments adhered to symptom-limited criteria and were terminated immediately if: (1) pain intensity exceeded pre-test baseline levels significantly; (2) a >20 mm increase on a 100-mm Visual Analog Scale (VAS) was reported (19); or (3) observable protective neuromuscular responses (e.g., guarding, withdrawal) were noted.

- Bilateral Manual Muscle Testing (MMT) Bilateral manual muscle testing (MMT) assessed isometric strength bilaterally for eight muscle groups critical to lumbopelvic stability: quadriceps, hamstrings, adductors, tensor fasciae latae, psoas major, piriformis, gluteus maximus, and gluteus medius. Testing followed standardized positions and protocols (20). For analytic purposes, strength outcomes were dichotomized to identify clinically relevant functional deficits. A score of “0” (Normal) was assigned if the participant maintained a strong isometric contraction against manual resistance. A score of “1” (Functional Deficit) was recorded if the contraction could not be sustained for ≥5 seconds (“break” phenomenon) or if marked bilateral asymmetry was observed. This binary coding was used to simplify model input and to identify clinically relevant functional deficits for logistic regression analysis.
- Assessment Lumbar Mobility Active lumbar range of motion was quantified using the dual-inclinometer technique. Two digital inclinometers (Hengshui Aohong Technology, China) were positioned over the T12 and S1 spinous processes. For flexion and extension, lumbar motion was estimated as the difference between the superior (T12) and inferior (S1) sensor readings to reduce pelvic contribution. Lateral side bending was measured using a single inclinometer at the T12 level, in accordance with established spinal flexibility assessment protocols.
- Hip Mobility Passive hip internal rotation (IR) and external rotation (ER) were measured bilaterally using a universal goniometer (Baseline®, Fabrication Enterprises Inc., USA). Participants were positioned prone with the knee flexed to 90°. To reduce pelvic compensation, a non-elastic strap was secured across the posterior superior iliac spines (PSIS). The examiner palpated the contralateral iliac crest throughout movement to monitor for pelvic rotation or tilt; trials showing compensatory motion were repeated (21, 22)
- Lumbopelvic-hip Functional Screening A battery of fourteen standardized orthopedic maneuvers was selected to evaluate LPH function (22, 23). Although traditionally used in differential diagnosis, these tests were operationally defined in the present study as functional screening assessments to identify mechanical sensitivity, mobility restriction, and aberrant motor control, rather than to establish specific pathoanatomical diagnoses. Tests were categorized into three domains: (1) neural sensitivity (e.g., slump test, straight leg raise); (2) articular loading and sensitivity (e.g., FABER test, quadrant test); and (3) myofascial extensibility (e.g., Thomas test, Ober’s test). For model development and interpretability, outcomes were dichotomized (0 = negative; 1 = positive for reproduction of familiar pain significant restriction, or observable compensatory movement patterns). A complete list of tests and operational definitions is provided in Supplementary Table S1.

### Establishment of the Clinical Reference Standard

In the absence of a singular pathological gold standard, a two-phase procedure involving expert consensus and reliability verification was used to establish the clinical reference standard before predictive modeling. In Phase 1, a multidisciplinary panel of seven senior clinicians (three dual-credentialed chiropractors/physical therapists, two ICCSP-certified sports chiropractors, and two licensed chiropractors; mean clinical experience: 12 years) defined structured triage criteria based on functional and orthopedic findings. Quantitative thresholds and decision rules were established a priori. In Phase 2, inter-rater reliability of the same predefined criteria was evaluated in a randomly selected subset of athletes (n = 41). Each panelist independently reviewed de-identified datasets comprising 53 variables and assigned triage classifications according to the prespecified rules, while remaining blinded to the classifications of the other raters. Agreement for the ordinal 4-level classifications was quantified using Fleiss’ weighted kappa, indicating high inter-rater reliability (*κ* = 0.977, 95% CI: 0.936–1.000) (24). The same criteria were then applied to the full analytic cohort (N = 864). Outcome classification for the analytic cohort was performed by a single calibrated examiner who had participated in the consensus process and applied the predefined criteria without modification. This approach was used to ensure consistent application of the predefined criteria across the full cohort. Participants were initially categorized into four triage grades reflecting clinical care needs: Grade 0 (healthy/asymptomatic), Grade 1 (low priority), Grade 2 (priority intervention), and Grade 3 (Red Flag). For predictive modeling, Grades 0 and 1 were combined as the self-management group (coded as 0), whereas Grade 2 was defined as the priority intervention group (coded as 1) (10, 25, 26). Athletes meeting Grade 3 criteria were referred for immediate physician evaluation and were excluded from predictive modeling. Thus, the reference standard reflected predefined stratification of intervention priority within an operational framework rather than structural diagnosis or long-term clinical outcome. Because this framework was based on symptom and functional findings, complete separation between predictors and outcome was not possible. However, model inputs consisted of individual raw symptom, functional, and examination variables rather than composite triage classifications, in order to reduce direct criterion contamination.

### Model Development & Validation

As prespecified, a complete-case analysis was performed. Missing data were not imputed because the proportion of incomplete cases was limited and the primary analysis was prespecified as complete-case analysis. Descriptive statistics were calculated for all demographic and clinical variables. Continuous variables were summarized as means and standard deviations (SDs), whereas categorical variables were summarized as frequencies and percentages. BMI was retained as a continuous variable during model development to preserve its linear risk gradient.

To construct a parsimonious model suitable for clinical translation for clinical translation, a forward stepwise multivariable logistic regression analysis was performed to identify independent variables associated with priority intervention. This approach was selected to derive a simplified and interpretable model that could be translated into an integer-based clinical triage score for field-based use. To reduce overfitting risk, candidate predictors were prespecified based on clinical reasoning, model complexity was evaluated using the events-per-variable (EPV) criterion, and internal validation was performed using bootstrap resampling. Candidate predictors entered into the model were prespecified based on clinical reasoning and included demographic characteristics, LPH functional assessment findings, and orthopedic provocation test results. Associations were expressed as odds ratios (ORs) with 95% confidence intervals (CIs). Sample size adequacy was evaluated using the EPV criterion, with a minimum threshold of 10 events per predictor considered indicative of acceptable model stability (27). Multicollinearity was assessed using variance inflation factors (VIFs), with values < 5 indicating no concerning collinearity. Overall model fit and performance were evaluated using the omnibus test, Nagelkerke *R*^*2*^, and the Brier score, with lower values indicating better overall accuracy (28). Internal validation was performed with 1,000 bootstrap resamples to assess model optimism and coefficient stability. Bootstrap-corrected estimates of the area under the receiver operating characteristic curve (AUROC) and calibration metrics were obtained, and bias-corrected and accelerated (BCa) 95% CIs were derived for regression coefficients (17, 18). Calibration was assessed using the Hosmer-Lemeshow test (*p* > 0.05 indicating acceptable fit). In addition, a calibration plot was generated to visually examine the alignment between observed and estimated probabilities along the ideal 45-degree reference line (28). The calibration slope was calculated by regressing observed outcomes on the logit-transformed predicted probabilities, with a slope of 1.0 indicating ideal calibration.

### Construction of the Weighted Clinical Triage Scoring System (CTSS)

To facilitate clinical implementation, the final multivariable logistic regression model was transformed into a weighted integer-based CTSS. Regression coefficients (β) were rescaled to preserve the relative contribution of each predictor while enabling straightforward point-based application (29). The absolute value of the smallest statistically significant coefficient in the final model was used as the scaling reference. A common scaling factor was derived from this reference, and each predictor was assigned an integer weight proportional to its relative effect size. For categorical predictors, integer weights were obtained by multiplying the corresponding β coefficients by the scaling factor and rounding to the nearest integer. For continuous predictors, such as BMI, weighted contributions were initially modeled on their original continuous scale to reflect their cumulative contribution across the observed clinical range. These were subsequently categorized into discrete intervals, as presented in Table 2, to improve interpretability and facilitate field-based use. An individual’s total CTSS score was calculated by summing the points assigned across all present predictors, with higher scores indicating greater priority for professional clinical evaluation and intervention.

**Table 2.**
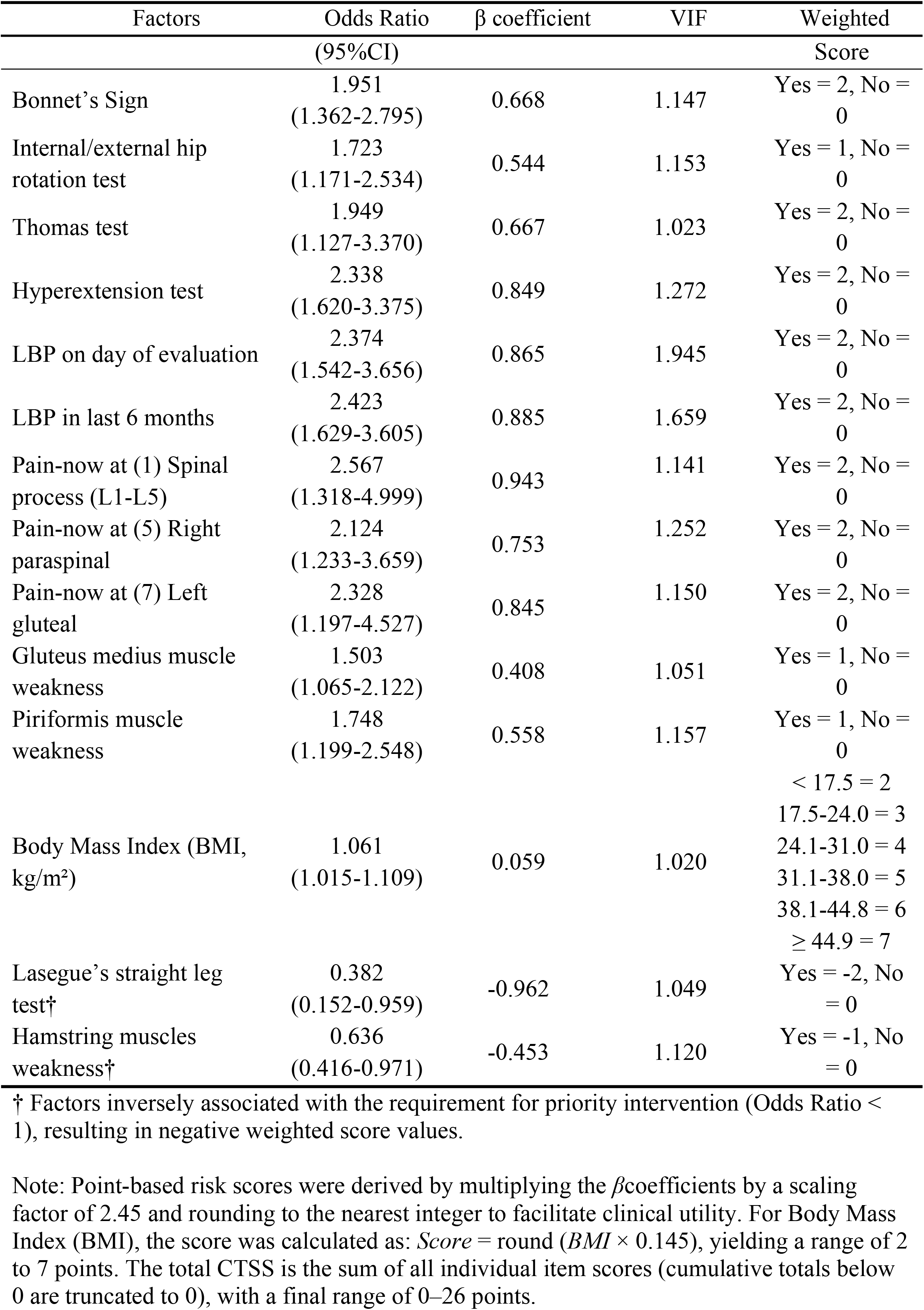
Multivariate Logistic Regression Model and Derived Weighted Scoring System for Identifying Athletes Requiring Priority Intervention.

### Evaluation and Clinical Utility Assessment

The performance of CTSS was evaluated in terms of discrimination, calibration, threshold-based classification, and clinical utility. Discriminative ability was assessed using the AUROC, reflecting the capacity of the scoring system to distinguish athletes requiring priority intervention from those suitable for self-management (17). Sensitivity, specificity, and overall classification accuracy were calculated across score thresholds. The optimal operating cutoff was determined using the Youden Index (sensitivity + specificity − 1) (30). To provide clinically interpretable risk estimates beyond dichotomous classification, the total CTSS score was also modeled as a continuous predictor within a logistic regression framework using the predefined binary reference standard as the dependent variable. Predicted probabilities of priority intervention were then derived for each possible score value, establishing a score– probability function for individualized probability estimation of priority intervention. Clinical utility was assessed using decision curve analysis (DCA) (31). Net benefit was estimated across a prespecified range of clinically relevant threshold probabilities (20%–50%), and the model was compared against default strategies of “treat-all” and “treat-none.” This threshold range was selected a priori to reflect realistic trade-offs between unnecessary referrals and missed priority cases in field-based sports medicine settings. All statistical analyses were performed using SPSS version 25.0 (IBM Corp., Armonk, NY, USA) with statistical significance set at *p* < 0.05. DCA was performed in Python 3.11.3.

## Results

### Demographic Characteristics and Clinical Classification

A total of 864 collegiate athletes, primarily from basketball, badminton, rugby, and taekwondo were included in the final analysis. Based on the predefined clinical reference standard for early rehabilitation triage, 463 athletes (53.6%) were classified into the priority intervention group and 401 (46.4%) into the self-management group. The cohort was predominantly male (n = 698, 80.8%) with a mean age of 21.3 ± 2.4 years. Participants reported an average sports participation duration of 9.7 ± 5.2 years and a weekly training volume of 20.4 ± 9.9 hours. On the day of assessment, the mean pain intensity was 3.25 ± 1.84 on the visual analogue scale.

The point prevalence of LBP was 47.7%, with 6-month and 12-month prevalence rates of 58.9% and 65.9%, respectively. A history of musculoskeletal injury was frequently reported, with the 3 most common sites being the ankle (58.9%), lower back (49.5%), and knee (40.7%). Regarding symptom distribution within the LPH zones, pain was most frequently identified in the left paraspinal (19.8%) and right paraspinal (18.4%) regions (Supplementary Figure S1). Detailed demographic characteristics and descriptive results of the LPH functional assessments, including range of motion, MMT, and orthopedic provocation tests, are presented in Table 1.

### Multivariable Logistic Regression and Internal Validation

Stepwise multivariable logistic regression identified 14 independent predictors from the predefined pool of 53 candidate variables (Table 2). With 463 priority intervention cases and 14 retained predictors, the EPV ratio was 33.1, exceeding the recommended minimum threshold of 10 and supporting model stability. The final model demonstrated satisfactory overall performance (Nagelkerke *R*^*2*^ = 0.470; omnibus test: *χ*^*2*^ = 374.515, *p* < 0.001; -2 log likelihood = 818.791). Multicollinearity was minimal, with VIF ranging from 1.020 to 1.945 (mean VIF = 1.224), indicating low collinearity among retained predictors. As shown in Table 2, factors positively associated with priority intervention included LBP within the previous 6 months, low back pain on the day of evaluation, localized pain at the L1–L5 spinous process, right paraspinal and left gluteal pain, positive hyperextension, Bonnet’s, Thomas, and internal/external rotation tests, gluteus medius weakness, piriformis weakness, and higher body mass index. In contrast, a positive Lasegue’s straight leg raise test and hamstring weakness were inversely associated with the outcome. Internal validation using 1,000 bootstrap resamples with replacement suggested good model stability. Bootstrap analysis showed that 12 of the 14 retained predictors (85.7%) remained statistically significant (p < 0.05), with one additional predictor showing marginal significance (hamstring weakness: bootstrap p = 0.077). All retained predictors demonstrated bootstrap 95% confidence intervals supporting their inclusion in the model. Calibration analysis also demonstrated good agreement between predicted and observed probabilities. Visual inspection of the calibration plot revealed close alignment with the 45° reference line (Figure 2A). The Hosmer–Lemeshow test indicated adequate goodness-of-fit (*χ*^*2*^ = 15.005, *p* = 0.059), and the Brier score was 0.152.

**Figure 2.**
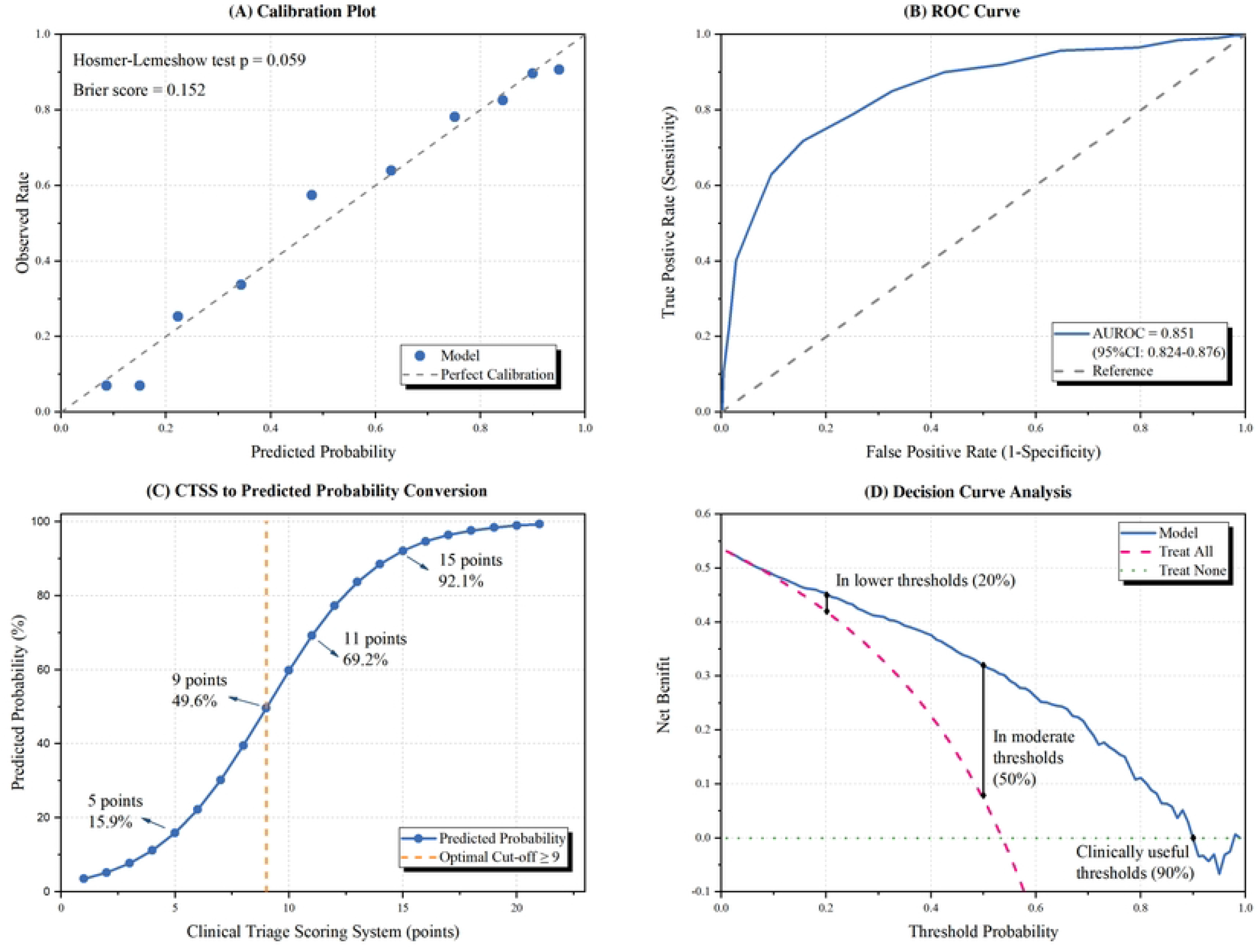
Predictive Performance and Clinical Utility of the Clinical Triage Scoring System (CTSS). (A) Calibration Plot: Observed vs. predicted probabilities showing close alignment with the 45° reference line (dashed). (B) ROC Curve: Discriminative performance of the CTSS. (C) Score-Probability Curve: Relationship between CTSS total score and predicted probability. Orange dashed line indicates optimal cut-off (≥ 9 points). (D) Decision Curve Analysis: Net benefit of the CTSS (blue line) compared to “Treat All” and “Treat None” strategies across threshold probabilities.

### Construction of the weighted CTSS

Regression coefficients from the final multivariable model were converted into an integer-based scoring system to derive the CTSS. Predictor weights were assigned by multiplying the *β* coefficients by 2.45 and rounding to the nearest integer. For BMI, which was retained as a continuous predictor, points were assigned using a graded scale to preserve usability in field-based scoring. The final CTSS ranged from 0 to 26 points, with higher scores indicating greater priority for further clinical evaluation and intervention (Table 2).

### Discrimination and threshold performance

The CTSS demonstrated good discrimination, with an AUROC of 0.851 (95% CI: 0.824– 0.876) (Figure 2B). The optimal threshold, determined by the maximum Youden index (0.563), was >8.5 points, corresponding to an operational threshold of ≥9 points (Table 3). At this threshold, sensitivity was 84.4% and specificity was 71.8%. At lower thresholds, such as ≥5 points, sensitivity increased to 97.2%, whereas specificity decreased to 40.1%. At higher thresholds, such as ≥11 points, specificity increased to 85.0%, whereas sensitivity decreased to 67.4%. These findings illustrate the trade-off between maximizing sensitivity and minimizing false-positive referrals across candidate triage thresholds.

**Table 3.** Coordinates of the ROC Curve showing Sensitivity, Specificity, and Youden Index at various CTSS cut-off values.

| Cut-off value | Sensitivity | Specificity | Youden index<br>(Sensitivity+ Specificity-1) |
| --- | --- | --- | --- |
| 0.0000 | 1.000 | 0.000 | 0.000 |
| 1.5000 | 0.998 | 0.000 | -0.002 |
| 2.5000 | 0.998 | 0.007 | 0.005 |
| 3.5000 | 0.996 | 0.107 | 0.103 |
| 4.5000 | 0.985 | 0.227 | 0.212 |
| 5.5000 | 0.972 | 0.401 | 0.373 |
| 6.5000 | 0.937 | 0.521 | 0.459 |
| 7.5000 | 0.905 | 0.628 | 0.533 |
| 8.5000 | 0.844 | 0.718 | 0.563 |
| 9.5000 | 0.749 | 0.788 | 0.537 |
| 10.5000 | 0.674 | 0.850 | 0.524 |
| 11.5000 | 0.575 | 0.900 | 0.475 |
| 12.5000 | 0.464 | 0.920 | 0.385 |
| 13.5000 | 0.352 | 0.958 | 0.310 |
| 14.5000 | 0.205 | 0.965 | 0.170 |
| 15.5000 | 0.127 | 0.985 | 0.112 |
| 16.5000 | 0.054 | 0.990 | 0.044 |
| 17.5000 | 0.028 | 0.995 | 0.023 |
| 18.5000 | 0.019 | 0.998 | 0.017 |
| 19.5000 | 0.004 | 0.998 | 0.002 |
| 20.5000 | 0.000 | 0.998 | -0.002 |
| 22.0000 | 0.000 | 1.000 | 0.000 |

Note: The optimal cut-off point was determined using the maximum Youden index (0.563),
corresponding to a score of $>8.5$ . For clinical application, this is operationalized as a threshold of $\geq 9$ points.

### Score-Probability Relationship

Logistic regression analysis demonstrated a positive association between CTSS score and classification into the priority intervention group. Each 1-point increase in CTSS score was associated with a 1.51-fold increase in the odds of priority intervention classification (OR = 1.510, 95% CI: 1.430–1.593). The derived score-probability relationship (Figure 2C) showed a graded, monotonic increase in the estimated probability of priority intervention classification across the observed score range. A CTSS score of 9 corresponded to an estimated probability of 49.6%.

### Clinical Utility

DCA was conducted to evaluate the clinical utility of CTSS (Figure 2D). Across the prespecified clinically relevant threshold probability range of 20%–50%, the model demonstrated consistently higher net benefit than both “treat-all” and “treat-none” strategies. At a 20% threshold probability, the CTSS yielded greater net benefit than the treat-all approach (0.454 vs. 0.420). This advantage persisted at the 50% threshold (0.319 vs. 0.072). These findings suggest that applying the CTSS within this probability range may improve referral efficiency by increasing the proportion of correctly identified priority cases while reducing unnecessary referrals.

## Discussion

The present study developed and internally validated the Clinical Triage Scoring System (CTSS), a 14-factor weighted scoring tool for field-based triage of lumbopelvic-hip (LPH) dysfunction in collegiate athletes. The CTSS was designed as a clinical decision-support tool for triage and care prioritization rather than as a diagnostic instrument or a predictive model for future injury. By integrating anthropometric, functional, and orthopedic provocation findings, it provides a structured framework for identifying athletes who may require priority intervention in field-based settings. In contrast to traditional clinical prediction rules targeting established LBP (9), the CTSS emphasizes the early recognition of movement-related and motor-control impairments relevant to athletic populations. The inclusion of LPH mobility and neuromuscular domains is consistent with prior evidence linking these factors to LBP in athletes (32). The CTSS demonstrated good discriminative performance (AUROC = 0.851), with an operational threshold of ≥9 points. Multicollinearity was minimal (VIF 1.02–1.95), and internal validation using bootstrap resampling suggested limited optimism and stable calibration. The small difference between the apparent and bootstrap-corrected AUROC was also consistent with the favorable events-per-variable ratio (33.1). When modeled as a continuous score, the CTSS showed a graded increase in the estimated probability of priority intervention classification, with a score of 9 corresponding to an approximate predicted probability of 50%. This supports its interpretation as a clinically meaningful decision boundary rather than an arbitrary dichotomous threshold. Although the CTSS comprises 14 items, its component variables are derived from routine clinical information and standardized field-based assessments, including BMI, pain history, range of motion, and MMT. This may support practical implementation in sports medicine settings without requiring advanced diagnostic hardware or complex software during initial triage. In the present cohort, 53.6% of athletes met the predefined criteria for priority intervention, including 19.5% who did not report spontaneous symptoms but demonstrated objective functional deficits. This finding suggests that the CTSS may identify clinically relevant impairment that is not evident from symptom reporting alone. Under the predefined reference standard, Grade 2 classification required multiple positive clinical findings in conjunction with objective impairment, a criterion intended to reduce over-classification based on isolated symptoms alone (15).

The retained predictors represent clinically relevant domains related to loading exposure, kinetic-chain function, and neuromuscular control. Higher BMI and a history of LBP may be associated with cumulative loading exposure and recurrent symptom burden, consistent with reports linking prior symptoms to recurrence risk (3). The inclusion of hip-centered markers, including Thomas test positivity, rotational mobility restrictions, and Bonnet’s sign, emphasizes the regional interdependence of the LPH complex rather than isolated lumbar symptoms (22, 23, 32, 33). Similarly, the pattern of paraspinal and gluteal findings, combined with hip muscle weakness, may reflect altered lumbopelvic coordination and compensatory loading patterns (5, 6, 34). The OR pattern further suggests that recent symptom burden (OR range 2.37–2.42), localized pain provocation, and clustered mechanical findings contributed more strongly to priority classification than isolated muscle weakness (OR range 1.50–1.75). BMI was the only continuous predictor retained and was converted into a graded point scale (2–7 points) to preserve its contribution while supporting usability in field-based scoring. The retention of multiple localized pain regions (L1–L5, paraspinal, and gluteal) further suggests that the CTSS may capture not only the presence of pain but also its topographical distribution, providing additional triage value beyond a binary history of LBP alone. Notably, the model assigned negative weights to a positive Lasegue’s sign (OR = 0.38) and hamstring weakness (OR = 0.64). Within this triage context, these findings may reflect posterior chain tightness or non-priority mechanical presentations rather than the specific functional profile captured by the CTSS. Because straight leg raise positivity in athletes may be influenced by musculotendinous stiffness and lumbopelvic movement characteristics rather than neural involvement alone, its isolated presence may be less informative for priority triage (35, 36). Thus, these negative weights may have helped reduce over-classification of athletes with non-specific flexibility limitations. However, any athlete demonstrating overt neurological deficits or progressive red-flag symptoms should undergo comprehensive medical evaluation.

An important strength of CTSS is its capacity to identify functional abnormalities in athletes who may not report prominent symptoms, thereby addressing a recognized limitation of symptom-based triage approaches (4). DCA further contextualized the clinical utility of the model (Figure 2D). Compared with treat-all and treat-none strategies, CTSS demonstrated consistently higher net benefit across the prespecified threshold probability range of 20%–50%. At the 50% threshold probability, corresponding to the operational score threshold of 9, the net benefit was 0.319, suggesting improved decision balance between identifying athletes requiring priority intervention and reducing unnecessary referrals. These findings suggest that structured triage may improve referral efficiency in field-based athletic settings, particularly where time and resources are constrained. However, CTSS is intended to support prioritization of further clinical evaluation and management, rather than to replace comprehensive clinical assessment, establish diagnosis, or make return-to-play decisions in isolation. Importantly, classification into the priority intervention group does not imply a single standardized treatment pathway. Rather, it indicates that the athlete may warrant timely professional assessment and targeted conservative management, while the specific treatment approach should remain individualized according to the full clinical presentation. Its application should be integrated with professional clinical judgment and appropriate medical evaluation when red flags or progressive symptoms are present. In this respect, CTSS aligns with broader clinical recommendations supporting early, targeted conservative care for LPH dysfunction when clinically appropriate (7).

Clinically, CTSS may serve as a structured decision-support framework for field-based triage. A score of ≥ 9 points identifies athletes with a higher probability of requiring priority intervention, whereas lower scores may support supervised self-management or continued monitoring, depending on the broader clinical context. The ≥9-point threshold should be interpreted as an operational threshold derived from the present cohort rather than a universally fixed cutoff. Notably, 19.5% of the priority-classified athletes reported no spontaneous symptoms at the time of assessment but were identified through objective functional findings, such as localized muscle weakness and positive provocation tests. This underscores the system’s capacity to detect clinically relevant impairments that are not captured by symptom reporting alone (37). By incorporating objective functional findings alongside symptom-related information, CTSS is consistent with rehabilitation approaches that emphasize movement quality and motor control in athletic populations (4). Nevertheless, the tool is intended to complement, rather than replace, comprehensive clinical evaluation, and its application should remain integrated with professional clinical judgment (8).

### Limitations

Several limitations should be acknowledged. First, the cross-sectional design captured current triage priority rather than longitudinal outcomes; thus, causal inference and prediction of future injury-related outcomes cannot be established. Second, the predominantly male (80.8%) cohort may limit generalizability to female athletes, professional or youth populations, and non-Asian groups (38). Given reported sex-related differences in lumbopelvic control, future studies may require model recalibration for broader application. Third, self-reported injury history may have introduced recall bias. Fourth, the expert-consensus reference standard did not include imaging; however, this was consistent with the study’s focus on functional triage rather than structural diagnosis, particularly given the documented weak correlation between imaging findings and clinical symptoms (39, 40). Although inter-rater agreement was high (kappa = 0.977), the reference standard remained consensus-based rather than an objective gold standard. Fifth, although conceptual overlap between the reference standard and candidate predictors could not be fully excluded, criterion contamination was partly reduced by entering individual raw clinical variables rather than composite triage grades into the model. Sixth, the stepwise regression approach has recognized limitations regarding variable selection stability. These risks were partly addressed through a favorable events-per-variable ratio (EPV = 33.1), bootstrap validation (n = 1,000) for optimism assessment, and low collinearity among retained predictors (VIF < 2.0). Finally, although the CTSS demonstrated good internal validity, external validation and formal evaluation of inter-rater reproducibility in real-world settings have not yet been conducted. Further studies across diverse athletic populations are warranted to assess generalizability, reproducibility, and clinical utility.

## Conclusion

In conclusion, this study developed and internally validated a 14-factor Clinical Triage Scoring System (CTSS) for the structured triage of LPH dysfunction in collegiate athletes. The model demonstrated good discrimination and satisfactory calibration, supporting its internal performance and stability as a clinical decision-support tool. Rather than functioning solely as a binary threshold-based tool, CTSS may also be interpreted on a continuous probability scale to support more individualized triage decisions in field-based athletic settings. The system is intended to complement, not replace, comprehensive clinical assessment. Future research should focus on external validation and on evaluating the impact of CTSS-guided triage on clinical resource allocation and athlete outcomes.

## Data Availability

All relevant data are within the manuscript and its Supporting Information files.

## Acknowledgments

We wish to thank all our participants for their time and effort. The study was supported by a grant from the NSYSU-KMU Joint Research Project (#NSYSUKMU 113-I01) and the National Science and Technology Council (NSTC) (#NSTC 113-2410-H-037-025-).

## References

1. Wilson F, Ardern CL, Hartvigsen J, Dane K, Trompeter K, Trease L, et al. Prevalence and risk factors for back pain in sports: a systematic review with meta-analysis. British Journal of Sports Medicine. 2021;55(11):601–7.

2. De Blaiser C, Roosen P, Willems T, Danneels L, Vanden Bossche L, De Ridder R. Is core stability a risk factor for lower extremity injuries in an athletic population? A systematic review. Physical therapy in sport. 2018;30:48–56.

3. Greene HS, Cholewicki J, Galloway MT, Nguyen CV, Radebold A. A history of low back injury is a risk factor for recurrent back injuries in varsity athletes. Am J Sports Med. 2001;29(6):795–800.

4. Hides JA, Jull GA, Richardson CA. Long-term effects of specific stabilizing exercises for first-episode low back pain. Spine (Phila Pa 1976). 2001;26(11):E243–E8.

5. Hungerford B, Gilleard W, Hodges P. Evidence of altered lumbopelvic muscle recruitment in the presence of sacroiliac joint pain. Spine (Phila Pa 1976). 2003;28(14):1593–600.

6. Vleeming A, Schuenke M, Danneels L, Willard F. The functional coupling of the deep abdominal and paraspinal muscles: the effects of simulated paraspinal muscle contraction on force transfer to the middle and posterior layer of the thoracolumbar fascia. J Anat. 2014;225(4):447–62.

7. Foster NE, Anema JR, Cherkin D, Chou R, Cohen SP, Gross DP, et al. Prevention and treatment of low back pain: evidence, challenges, and promising directions. Lancet. 2018;391(10137):2368–83.

8. Azevedo VD, Ferreira Silva RM, de Carvalho Borges SC, Fernades MdSV, Miñana-Signes V, Monfort-Pañego M, et al. Instruments for assessing back pain in athletes: a systematic review. PLoS One. 2023;18(11):e0293333.

9. Flynn T, Fritz J, Whitman J, Wainner R, Magel J, Rendeiro D, et al. A clinical prediction rule for classifying patients with low back pain who demonstrate short-term improvement with spinal manipulation. Spine (Phila Pa 1976). 2002;27(24):2835–43.

10. Alrwaily M, Timko M, Schneider M, Stevans J, Bise C, Hariharan K, et al. Treatment-based classification system for low back pain: revision and update. Phys Ther. 2016;96(7):1057–66.

11. Bahr R. Why screening tests to predict injury do not work—and probably never will…: a critical review. British journal of sports medicine. 2016;50(13):776–80.

12. Macmahon B, Weiss NS, Pocock SJ, Gøtzsche PC, Vandenbroucke JP, Kuller LH. The strengthening the reporting of observational studies in epidemiology (STROBE) statement: guidelines for reporting observational studies. Commentary: strobe initiative. Epidemiology (Cambridge, Mass). 2007;18(6).

13. Delitto A, George SZ, Van Dillen L, Whitman JM, Sowa G, Shekelle P, et al. Low back pain: clinical practice guidelines linked to the International Classification of Functioning, Disability, and Health from the Orthopaedic Section of the American Physical Therapy Association. J Orthop Sports Phys Ther. 2012;42(4):A1–A57.

14. Dionne CE, Dunn KM, Croft PR, Nachemson AL, Buchbinder R, Walker BF, et al. A consensus approach toward the standardization of back pain definitions for use in prevalence studies. Spine (Phila Pa 1976). 2008;33(1):95–103.

15. Bahr R, Clarsen B, Derman W, Dvorak J, Emery CA, Finch CF, et al. International Olympic Committee consensus statement: methods for recording and reporting of epidemiological data on injury and illness in sports 2020 (including the STROBE extension for sports injury and illness surveillance (STROBE-SIIS)). Orthop J Sports Med. 2020;8(2):2325967120902908.

16. Clarsen B, Myklebust G, Bahr R. Development and validation of a new method for the registration of overuse injuries in sports injury epidemiology: the Oslo Sports Trauma Research Centre (OSTRC) overuse injury questionnaire. Br J Sports Med. 2013;47(8):495–502.

17. Collins GS, Reitsma JB, Altman DG, Moons KGM. Transparent reporting of a multivariable prediction model for individual prognosis or diagnosis (TRIPOD): the TRIPOD statement. Br J Surg. 2015;102(3):148–58.

18. Steyerberg EW, Harrell FE Jr, Borsboom GJ, Eijkemans M, Vergouwe Y, Habbema JDF. Internal validation of predictive models: efficiency of some procedures for logistic regression analysis. J Clin Epidemiol. 2001;54(8):774–81.

19. Childs JD, Piva SR, Fritz JM. Responsiveness of the numeric pain rating scale in patients with low back pain. Spine (Phila Pa 1976). 2005;30(11):1331–4.

20. Conable KM, Rosner AL. A narrative review of manual muscle testing and implications for muscle testing research. J Chiropr Med. 2011;10(3):157–65.

21. Han S, Kim RS, Harris JD, Noble PC. The envelope of active hip motion in different sporting, recreational, and daily-living activities: a systematic review. Gait Posture. 2019;71:227–33.

22. Calcei JG, Safran MR. Evaluation of athletes with hip pain. Clin Sports Med. 2021;40(2):221–40.

23. Peebles R, Jonas CE. Sacroiliac joint dysfunction in the athlete: diagnosis and management. Curr Sports Med Rep. 2017;16(5):336–42.

24. McHugh ML. Interrater reliability: the kappa statistic. Biochemia medica. 2012;22(3):276–82.

25. Qaseem A, Wilt TJ, McLean RM, Forciea MA; Clinical Guidelines Committee of the American College of Physicians. Noninvasive treatments for acute, subacute, and chronic low back pain: a clinical practice guideline from the American College of Physicians. Ann Intern Med. 2017;166(7):514–30.

26. Bastos RM, Moya CR, de Vasconcelos RA, Costa LOP. Treatment-based classification for low back pain: systematic review with meta-analysis. J Man Manip Ther. 2022;30(4):207–27.

27. Peduzzi P, Concato J, Kemper E, Holford TR, Feinstein AR. A simulation study of the number of events per variable in logistic regression analysis. J Clin Epidemiol. 1996;49(12):1373–9.

28. Van Calster B, Nieboer D, Vergouwe Y, De Cock B, Pencina MJ, Steyerberg EW. A calibration hierarchy for risk models was defined: from utopia to empirical data. J Clin Epidemiol. 2016;74:167–76.

29. Sullivan LM, Massaro JM, D’Agostino RB Sr. Presentation of multivariate data for clinical use: the Framingham Study risk score functions. Stat Med. 2004;23(10):1631–60.

30. Youden WJ. Index for rating diagnostic tests. Cancer. 1950;3(1):32–5.

31. Vickers AJ, Holland F. Decision curve analysis to evaluate the clinical benefit of prediction models. Spine J. 2021;21(10):1643–8.

32. Harris-Hayes M, Sahrmann SA, Van Dillen LR. Relationship between the hip and low back pain in athletes who participate in rotation-related sports. J Sport Rehabil. 2009;18(1):60–75.

33. Van Dillen LR, Bloom NJ, Gombatto SP, Susco TM. Hip rotation range of motion in people with and without low back pain who participate in rotation-related sports. Phys Ther Sport. 2008;9(2):72–81.

34. Sebyani M, Minoonejad H, Shirzad E, Alizadeh MH, Sarvestan J. Electromyography activity of posterior oblique sling muscles during high-speed running in elite soccer players: risk factors for hamstring injury. BMC Sports Sci Med Rehabil. 2025;18(1):74.

35. Miyamoto N, Hirata K, Kimura N, Miyamoto-Mikami E. Contributions of hamstring stiffness to straight-leg-raise and sit-and-reach test scores. Int J Sports Med. 2018;39(02):110–4.

36. Santonja-Medina F, Santonja-Renedo S, Cejudo A, Ayala F, Ferrer V, Pastor A, et al. Straight leg raise test: influence of Lumbosant© and assistant examiner in hip, pelvis tilt and lumbar lordosis. Symmetry. 2020;12(6):927.

37. Fett D, Trompeter K, Platen P. Back pain in elite sports: a cross-sectional study on 1114 athletes. PLoS One. 2017;12(6):e0180130.

38. Hart DA. Sex differences in musculoskeletal injury and disease risks across the lifespan: are there unique subsets of females at higher risk than males for these conditions at distinct stages of the life cycle? Front Physiol. 2023;14:1127689.

39. Brinjikji W, Luetmer PH, Comstock B, Bresnahan BW, Chen L, Deyo R, et al. Systematic literature review of imaging features of spinal degeneration in asymptomatic populations. AJNR Am J Neuroradiol. 2015;36(4):811–6.

40. Maher C, Underwood M, Buchbinder R. Non-specific low back pain. Lancet. 2017;389(10070):736–47.

